# Community-Based Hypertension Screening in Urban and Rural Zimbabwe: A Comparative Analysis and Implementation Lessons (2022–2025)

**DOI:** 10.64898/2026.11.23.26368728

**Authors:** Pugie T. Chimberengwa, Kenneth Masiye, Tafadzwa O. Bepe, Trudy T. Mhlanga, Efison Dhodho, Tinashe T. Chinyanga

**Affiliations:** Organization for Public Health Interventions and Development, Harare, Zimbabwe; Department of Community Medicine, Faculty of Medicine, National University of Science and Technology, Bulawayo, Zimbabwe

**Keywords:** Hypertension, Community health workers, Community-based screening, Primary healthcare, Decentralized care, Zimbabwe

## Abstract

**Introduction:** Hypertension is a leading cause of cardiovascular disease and a growing public health challenge in sub-Saharan Africa. Community-based screening delivered through community health workers (CHWs) may improve early detection and strengthen decentralized chronic disease services, particularly in underserved settings. This study compared hypertension screening yield across phased implementation of a community-based programme in urban and rural Zimbabwe and documented implementation lessons for decentralized hypertension services.

**Methods:** A retrospective comparative analysis was conducted using aggregate programme data routinely captured in the District Health Information System 2. Screening outcomes from Phase 1 (2022-2023) were compared with those from the combined Phase 2 and Phase 3 implementation period (2024-2025). Trained CHWs used digital blood pressure devices to screen adults in communities and outreach settings, refer individuals with elevated blood pressure to primary healthcare facilities, and support follow-up. Phase 1 was implemented in urban districts and targeted adults aged ≥40 years, while subsequent phases expanded to adults aged ≥18 years and predominantly rural and peri-urban populations. The primary outcome was hypertension screening positivity, while secondary outcomes included participant characteristics and age-specific positivity rates.

**Results:** A total of 410,568 adults were screened, including 120,144 urban adults aged ≥40 years in Phase 1 and 290,424 adults in Phases 2 and 3. Among participants in Phases 2 and 3, 76.0% resided in rural communities, 64.9% were female, and 44.9% were living with HIV. Hypertension positivity was 41.0% (49,276/120,144; 95% CI 40.7%-41.3%) in Phase 1 and 14.5% (41,972/290,424; 95% CI 14.3%-14.6%) in Phases 2 and 3 (p<0.001). Positivity increased significantly with age in both periods, ranging from 31.8% to 51.6% in Phase 1 and from 13.5% to 28.4% in later phases (all p<0.001).

**Conclusion:** Community-based hypertension screening delivered by CHWs successfully reached large urban and rural populations and identified substantial hypertension burden, particularly among older adults. The lower positivity observed in later phases likely reflects differences in age eligibility and population mix rather than programme effect. Scaling CHW-led screening may strengthen decentralized hypertension detection, although nationally representative surveys and longitudinal studies are needed to assess linkage to care, treatment initiation, and long-term blood pressure control.

**What is already known on this topic:** Hypertension is a leading cause of cardiovascular disease in sub-Saharan Africa, but many people remain undiagnosed, especially in rural and underserved communities. Community health worker (CHW)-led screening has been shown to be a feasible approach for increasing access to hypertension detection and improving case finding beyond health facilities. However, evidence from large-scale routine programme implementation is limited, particularly in Southern Africa. There is also limited understanding of how screening yield varies across urban and rural settings during programme scale-up. This study was conducted to evaluate hypertension screening outcomes and implementation experiences from a large CHW-led programme in Zimbabwe.

**What this study adds:** This study provides evidence from one of the largest community-based hypertension screening programmes implemented through CHWs in Zimbabwe. It demonstrates that CHW-led screening can successfully reach large urban and rural populations and identify substantial numbers of people with elevated blood pressure. The findings show that hypertension positivity increases markedly with age and remains particularly high among older adults. The study also highlights important programme implementation lessons from scaling services across different settings and shows that differences in screening yield are strongly influenced by population characteristics, especially age distribution and rural-urban context, rather than programme phase alone.

**How this study might affect research, practice or policy:** The findings support the integration of CHW-led hypertension screening into routine primary healthcare and community outreach services as a strategy for expanding early detection of hypertension in Zimbabwe and similar settings. The results highlight the importance of prioritizing older adults, who had the highest screening positivity, while maintaining broad community access. Policymakers may use this evidence to strengthen investment in decentralized non-communicable disease services, digital screening tools, and CHW training. Future research should evaluate linkage to care, treatment initiation, retention in care, and blood pressure control outcomes to determine the long-term effectiveness and impact of community-based hypertension screening programmes.

## Introduction

Low- and middle-income countries (LMICs) are facing a dual burden of communicable and non-communicable diseases (NCDs). Premature mortality from NCDs is modelled to be 33% by 2030 [1,2]. Research has shown that hypertension (HT) commonly co-exists with chronic kidney disease, diabetes mellitus (DM), and hypercholesterolemia [3]. Recipients of care (RoC) in LMICs with NCDs had poor outcomes along with limited prospects for a healthy life due to poor access to healthcare services. Diabetes mellitus and HT are major contributors to global morbidity and premature mortality, with rapid growth in LMICs [2,4,5]. Thus, lowering blood pressure is a priority in individuals with DM to prevent cardiovascular complications [6].

The intersection between NCDs and communicable diseases, especially human immunodeficiency virus (HIV) further complicates this landscape with people living with HIV (PLHIV) increasingly affected by cardiometabolic disorders due to prolonged life expectancy from antiretroviral therapy (ART) use [7]. Comorbid disease management also loosely referred as multimorbidity disease management is becoming increasingly important in the long-term management of recipients of care (RoC) who are stable on ART [8].

Hypertension is defined as a persistent systolic blood pressure (BP) reading of 140 mm Hg or more and/or a diastolic BP of more than 90 mm Hg [1,2,9,10]. Globally less than 20% of people living with hypertension (PLHT) have controlled blood pressure [5,11,12] Thus, due to their chronicity nature, NCDs strain the already scarce resources of healthcare systems and families in LMICs [13]. While substantial concern for NCDs centers around the impact of urbanization in sub-Saharan Africa (sSA), the growing burden of HT in rural areas remains largely neglected. However, the increase in the burden of NCDs in rural LMICs is entirely not related to sedentary lifestyles and neither high calory diets [13].

Globally, it is estimated that 1.13 billion people are living with HT, mostly in LMICs and the highest prevalence recorded in sSA is associated with poor awareness and control [1,6,10]. In Africa, the World Health Organization (WHO) estimates that, 46% of adults aged 25+ years, were living with HT compared to 35% -40% in other parts of the world [1,3,14,15]. Most Africans living with HT are unaware of their status and are less likely to seek treatment or are poorly controlled, predisposing them to high risk for cardiovascular diseases (CVDs) [1,16]. There is a paucity of national data on HT prevalence studies in Zimbabwe, however, the STEPwise survey in 2005, reported the national HT prevalence at 27% [17].

Hypertension is the major risk factor for global cardiovascular morbidity and mortality, with about half of cardiovascular events attributed to HT, [5,12,18] and that significantly increases the risks of developing CVDs [16,19,20]. Randomized control studies have shown that that strict BP control in RoCs with HT reduces the risk and related complications such as heart failure, and death [6]. Lowering HT reduces the risk by 50% for heart failure, myocardial infarction by 20–25%, and stroke by 35–40% [14]. Hypertension can easily be screened and diagnosed in the community and primary care (PHC) facilities, and governments can enable access to affordable and effective medicines [5,15,21]. In African communities, the challenges in managing HT lie in prevention, diagnosis and treatment [22]. However, elevated BP is generally asymptomatic from onset to several years, and at diagnosis, RoC may have developed complications which results in high morbidity and mortality [23].

The NCDs integration project was implemented by the Organization for Public Health Interventions and Development (OPHID) in support of the Ministry of Health and Child Care (MOHCC) in Zimbabwe with catalytic support from Sanofi. The project was implemented in phases starting with urban areas (Bulawayo and Chitungwiza - phase 1) in September 2022, then the subsequent phases were expanded in the rural areas (Gwanda, Insiza, Beitbridge and Umzingwane - phase 2 and 3). Phase 3 was completed in September 2026, and the project was handed over to MOHCC. The aim of the project was to integrate hypertension and diabetes mellitus screening, diagnosis, treatment and control in PLHIV and the general population. This then formed a basis for this study which was part of an ongoing public health program in the MOHCC, Zimbabwe.

## Methods

We conducted a retrospective, comparative analysis of aggregate programme screening records extracted from the District Health Information System version 2 (DHIS2) comparing results for Hypertension screening from Phase 1 implementation (2022–2023) and combined Phase 2 and 3 (2024–2025).

The NCDs program implementation was in a phased approach. Phase 1 was implemented in two urban areas of Bulawayo and Chitungwiza in Zimbabwe from September 2022 to March 2024. In this phase adults aged 40 years and above were provided with HT screening, diagnosis, treatment and follow-up as part of the program. Phase 2 was expanded to adults aged 18 years and above in the same districts in phase 1 and additional two districts in Matebeleland South province namely Gwanda predominantly rural (but has both rural and urban) and Insiza which is a rural district though unique in that there is a high population of artisanal miners. In phase 3, a border town with both rural and urban population (Beitbridge) and peri-urban rural district of Umzingwane were added.

Hypertension screening services were predominantly done by trained community health workers, community outreach agents (COAs) who were expert clients living positively with HIV. The COAs were leveraged from the Target Accelerate and Sustain Quality Care for HIV Epidemic Control (TASQC) program that OPHID was implementing in these and other districts in support of the MOHCC. The COAs were trained to record and interpret BP readings using battery operated digital BP machines. The program procured these machines for their use within the communities. Whenever they found a RoC with elevated BP, they would refer them to the nearest health facility as part of the linkage process or contact the TASQC program supported nurses to follow up the clients in the community. A follow up of the clients with HT was done to check if they were linked to public health facilities for care, treatment and follow up.

Data was anonymized and coded for ease of analysis and to remove individualized patient details to ensure data privacy. Descriptive statistics were used to summarize demographic characteristics, HIV status, service delivery geography (urban or rural residence), and hypertension screening outcomes. Categorical variables were summarized using frequencies and percentages, with percentages calculated using the relevant phase-group denominators. Phase 1 data were pooled across 2022 and 2023 to provide the urban baseline, while Phase 2 and 3 data were combined across 2024 and 2025. Hypertension screening yield was summarized using the number of participants who screened positive, the number screened, and the corresponding percentage. Pearson chi-square tests were used to compare categorical distributions and HT positivity between Phase 1 and combined Phases 2 and 3. Participant characteristics were compared by sex, HIV status, residence, and age group. Age-specific HT positivity was also compared between phase groups among adults aged 40 years and above, using the age bands 40–44, 45–49, 50–54, 55–59, 60–64, and 65 years and above. Comparisons between phases were interpreted considering differences in the age and urban–rural composition of the screened populations.

## Results

A total of 120,144 participants were screened in Phase 1 and 290,424 in the combined Phase 2 and 3. Phase 1 was for an urban population aged 40 years and above in Bulawayo and Chitungwiza. Phases 2 and 3 recorded more rural population (76%) compared to the urban population. Those below the age of 40 were 52% compared to those aged 40 years and above.

**Table 1.** Participant characteristics in Phase 1 versus combined Phase 2& 3.

| Variable | Category | Phase 1; n (%) | Phase 2&3 combined; n (%) | p-value |
|---|---|---|---|---|
| **Sex** | Male | 44,747 (37.2%) | 101,921 (35.1%) | <0.001 |
|  | Female | 75,397 (62.8%) | 188,503 (64.9%) | <0.001 |
| **HIV status** | HIV-negative | 71,005 (59.1%) | 160,095 (55.1%) | <0.001 |
|  | HIV-positive | 49,139 (40.9%) | 130,329 (44.9%) | <0.001 |
| **Residence** | Urban | 120,144 (100.0%) | 69,591 (24.0%) | <0.001 |
|  | Rural | 0 (0.0%) | 220,833 (76.0%) | <0.001 |
| **Age group** | ≥40 years | 120,144 (100.0%) | 139,295 (48.0%) | <0.001 |
|  | 18-40 years | 0 (0.0%) | 151,129 (52.0%) | <0.001 |

Most RoCs were female in both phases (62.8% and 64.9%) respectively. The sex distribution differed modestly between the two groups, with men accounting for 37.2% of participants in Phase 1 and 35.1% in combined Phase 2 and 3 (p<0.001). There were more HIV negative RoCs screened 59.1% and 55.1% in phase 1 and combined phase 2 &3 respectively. The HIV status distribution also differed significantly: participants documented as HIV-positive comprised 40.9% of Phase 1 compared with 44.9% in combined Phase 2 and 3 (p<0.001).

**Table 2.**
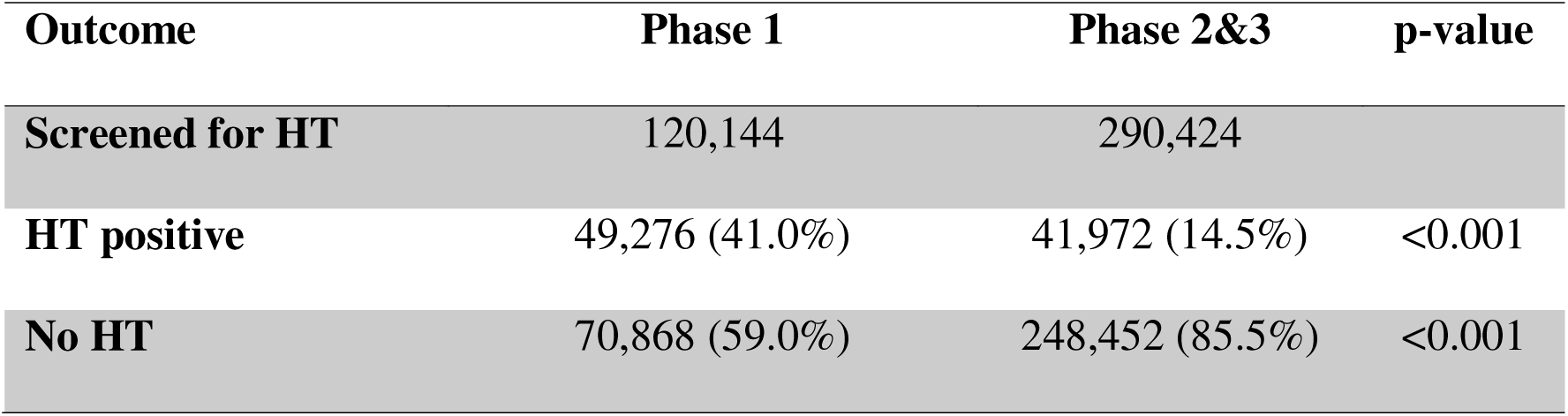
Hypertension screening outcomes in Phase 1 versus combined Phase 2&3.

Phase 1 showed a markedly higher recorded HT positivity rate (41%) than the pooled later phases (14.5%). However, this difference should not be interpreted as a pure phase effect because the comparison is confounded by major structural shifts in the screened population, especially the addition of younger adults and rural participants in the later years.

The prevalence of HT increased significantly after the age of 40 years and was higher in the urban compared to rural populations.

**Table 3.**
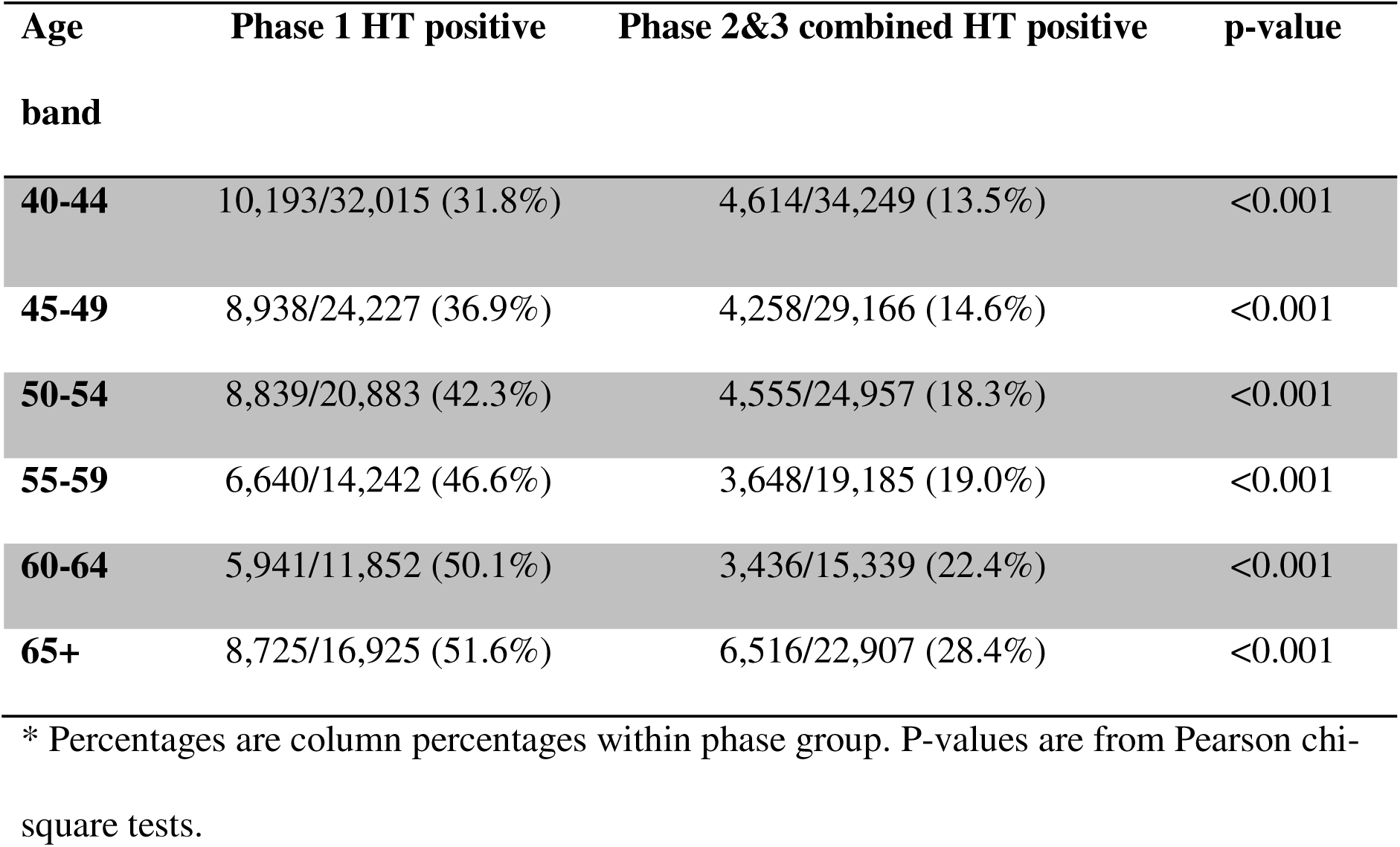
Age-specific hypertension positivity among adults aged 40 years and above.

| **Age band** | **Phase 1 HT positive** | **Phase 2&3 combined HT positive** | **p-value** |
|---|---|---|---|
| **40-44** | 10,193/32,015 (31.8%) | 4,614/34,249 (13.5%) | <0.001 |
| **45-49** | 8,938/24,227 (36.9%) | 4,258/29,166 (14.6%) | <0.001 |
| **50-54** | 8,839/20,883 (42.3%) | 4,555/24,957 (18.3%) | <0.001 |
| **55-59** | 6,640/14,242 (46.6%) | 3,648/19,185 (19.0%) | <0.001 |
| **60-64** | 5,941/11,852 (50.1%) | 3,436/15,339 (22.4%) | <0.001 |
| **65+** | 8,725/16,925 (51.6%) | 6,516/22,907 (28.4%) | <0.001 |

\* Percentages are column percentages within phase group. P-values are from Pearson chi-square tests.

## Discussion

During Phase 1, the programme focused on adults aged 40 years and older to establish baseline data on HT prevalence among a high-risk population. This approach was informed by the absence of recent national data, as Zimbabwe’s most recent WHO STEPS survey had been conducted in 2005. Given the catalytic nature of the funding, programme resources were strategically targeted to a population most likely to benefit from HT screening. The findings revealed a HT positivity rate of 41%, highlighting a substantial burden of disease and an important public health concern. However, this prevalence was consistent with reports from other studies conducted across sSA. These findings subsequently generated interest in understanding the burden of HT among younger adults and populations residing in rural areas, leading to the program expansion in later phases.

A systematic review on the burden of undiagnosed HT in sSA reported HT prevalence ranging between 15% and 70%, with a pooled prevalence of 30% (27–34%) [2]. These findings are comparable to our study findings where pooled prevalence was 14.5% including the higher prevalence of HT in those above the age of 40 years. In keeping with the high prevalence of HT in sSA, Zimbabwe was not spared. Similarly, the standstill in global prevalence and the global HT control rates of approximately 20% should serve as an important global wakeup call that CVDs are going to be a burden for many years to come, especially if we do not pay particular attention to influence change [7].

In our study, HT prevalence increased progressively with age, from 40 years to ≥65 years, irrespective of whether participants resided in urban or rural settings. However, across all 10-year age bands above 40 years, urban residents were approximately twice as likely to have HT as their rural counterparts. This disparity may reflect differences in lifestyle and environmental exposures, including lower levels of physical activity, more sedentary occupations, and greater consumption of unhealthy diets among urban populations. Previous studies have identified several risk factors that contribute to the high burden of hypertension in urban settings. In LMICs, the growing prevalence of HT has been strongly associated with four key behavioral risk factors: physical inactivity, unhealthy diet, tobacco use, and harmful alcohol consumption [24].

During Phase 1, all participants were recruited from urban areas. As the programme expanded into rural districts during subsequent phases, a substantially larger number of individuals were screened for HT in rural communities. The screening data suggest that rural populations may have been more effectively reached by the programme than their urban counterparts. A possible explanation is that COAs were predominantly based in rural areas, where they conducted household-level screening, whereas in urban settings screening was largely conducted through periodic outreach activities. Population-based HT screening, effective patient follow-up, and strong community-based primary healthcare systems are recognized as key strategies for the control of HT [10]. The program’s community-centered approach therefore appears to have contributed to increased screening coverage and the identification of large numbers of individuals with elevated BP, particularly in underserved rural areas.

In our study, 59% of individuals screened for HT were HIV-negative, while a smaller proportion were PLHIV. Given that the national HIV prevalence in Zimbabwe was approximately 12% in 2025, this finding suggests that PLHIV may have better health-seeking behavior and greater engagement with health services than HIV-negative individuals. As HIV and NCDs increasingly coexist, they can no longer be managed as separate conditions, particularly in high HIV-burden settings [2]. These findings also underscore the need to strengthen NCDs screening and preventive health-seeking behavior among the general HIV-negative population, especially men. Although DM was not a focus of this study, disorders of glucose metabolism are common among PLHIV, with diabetes prevalence estimated at around 10% among individuals receiving antiretroviral therapy (ART), compared with 3% among ART-naïve populations [8]. HIV-related factors, including chronic inflammation, lipodystrophy, comorbidities, and ART exposure, may contribute to the development of diabetes at lower levels of adiposity than in people without HIV [8].

There were considerable knowledge gaps related to the health and economic cost of HT and CVDs in LMICs [25]. The benefits of tight HT control in patients with diabetes exceed the benefits of tight glycemic control and extend to the prevention of macrovascular and microvascular complications [9]. Compliance with treatment of HT and DM has various benefits for the individuals, the health care systems and the society at large in the case of cost saving since it reduces the incidence of complications and the need for additional medications [6,9].

In this study there were 48% adults above the age of 40 years who were screened for HT compared to those below 40 years despite that there were more younger people under the age of 40 in the general population. This speaks to the health seeking behavior of adults who were more health conscious to the development of NCDs. On a similar note, there were more females (63%) than men, this trend is similar for both urban and rural areas. It has traditionally been the case across the healthcare system with men having poor health seeking behavior.

Blood pressure is strongly influenced by age. Consistent with previous evidence, our study found that HT prevalence increased with age, with the highest rates observed among adults aged 40 years and older. Studies have demonstrated a graded relationship between BP and CVD risk, even at levels as low as 115/75 mmHg [1], and systolic BP is known to rise progressively after 30-40 years of age before plateauing in later life [18]. Although fewer younger adults were screened, the findings revealed a notable burden of HT among those under 40 years. This highlights the need to challenge misconceptions that HT is a disease of older age and to strengthen public health education and screening initiatives targeting all adult age groups [18].

During programme scale-up, several implementation challenges were identified and addressed. As Phase 1 was a pilot, it encountered initial operational constraints. Although HT screening is traditionally conducted at PHC facilities in Zimbabwe, the programme introduced community-based screening and deployed BP devices to community health workers. This revealed gaps in service readiness, as some PHC facilities lacked functional BP machines for confirming diagnoses. In response, additional equipment was provided to strengthen facility capacity. Furthermore, when individuals with elevated BP were referred to PHC facilities, some healthcare workers lacked confidence in HT screening, diagnosis, and management. Targeted coaching, mentorship, and capacity-building were subsequently provided, improving service delivery and strengthening the continuum of HT care.

Access to physicians and antihypertensive medicines posed significant challenges during programme implementation. In urban areas, doctors were available at PHC facilities only on designated days, while in rural areas consultations were often limited to irregular outreach visits. Consequently, some clients incurred repeated consultation and travel costs before receiving a confirmed diagnosis. Medicine shortages at PHC facilities further affected continuity of care, forcing patients to seek treatment from higher-level facilities or private pharmacies. These barriers negatively impacted linkage to care. To address them, the programme integrated doctors into outreach activities, partnered with private pharmacies to provide free medications during selected outreaches, and promoted pooled community procurement mechanisms to improve access to essential HT medicines.

Access to HT care remains limited in many sSA settings. Studies from South Africa have shown that fewer than 5% of patients in rural sSA have access to appropriate HT care [13]. In Zimbabwe, access to NCD medicines at PHC facilities has been constrained by weak forecasting and quantification systems, partly due to limited HT surveillance data. To address this gap, the programme routinely shared screening data with the MOHCC NCDs department to support forecasting, quantification, and planning. However, the absence of longitudinal patient tracking limited the ability to assess linkage to care, treatment adherence, and BP control among screened individuals.

Improving adherence is particularly important because HT is asymptomatic and long-term treatment adherence remains poor [1]. Non-adherence is a major contributor to uncontrolled HT [2]. Understanding barriers to BP control in sSA can help policymakers prioritize interventions, guide health system strengthening, and identify unmet patient needs [12]. Digital health strategies, particularly mobile health (m-Health) interventions for patient education and follow-up, offer promising opportunities to improve hypertension management [1].

Despite significant research and policy efforts, global progress in HT control remains slow, highlighting the need for innovative approaches that strengthen diagnosis, treatment, and long-term management through primary care and community-based systems [7]. Opportunistic screening is a practical strategy for identifying individuals unaware of their BP status and linking them to care [1]. In line with the WHO Package of Essential Noncommunicable (PEN) Disease Interventions for PHC in Low-Resource Settings, training, and capacity building of community health workers and PHC clinicians are critical to improving HT outcomes [1]. While HT can be effectively managed at PHC level through lifestyle modification, counselling, and medication, many health systems in low-income settings remain inadequately equipped to provide sustained care [24]. Socioeconomic inequalities further influence HT burden and access to care, underscoring the need for equitable, patient-centred approaches to lifelong HT management [9,26].

## Limitations

This study was a desk review of program data; therefore, sub-analysis of some indicators could not be properly done as this could not have been the primary reason for program implementation. The lessons learnt were drawn from the implemented program and were not part of the objectives of data analysis and thus could not be reported in the results though they were key to explaining some of the data in the results as shared in the discussion. There was no longitudinal tracking for individual clients and thus indicators on linkage and control of HT were not precisely recorded in the study. The sampled districts were not representative of the general population as they were part of implementation district in the TASQC program and thus the findings cannot be generalized for the Zimbabwean population. However, the same findings can be used for inference and as a baseline data for a healthcare system that has not done a nationwide HT prevalence survey since 2005.

We therefore recommend more focused research on generating national baseline surveys on the prevalence of HT. As well there should be longitudinal tracking of RoC to include linkage to care and control of HT in Zimbabwe. These efforts should contribute to the solving the gaps that are there in NCDs care in Zimbabwe and the SSA.

## Conclusion

This study contributes to the body of knowledge in the Zimbabwe NCDs discourse. The findings confirm the high prevalence of HT in Zimbabwe mainly related to a sedentary lifestyle in the urban population. It also highlights the need for the younger generation to regularly screen for HT as there seems to be a recorded risk in this population. The PLHIV who regularly visit PHC for services should be educated to seek NCDs screening. Access to HT and NCDs screening in general should be decentralized within the communities though CHW screening programs and outreach activities if the country wants to be ahead in tackling the growing burden on NCDs.

## Data Availability

All data produced in the present study are available upon reasonable request to the authors

## Ethical Approval

Not applicable

## Author Contributions

**Pugie Chimberengwa**

Roles: Conceptualization, Supervision, Writing – Original Draft Preparation, Funding Acquisition, Writing – Review & Editing

**Tafadzwa Bepe**

Roles: Investigation, Methodology, Writing – Original Draft Preparation

**Kenneth Masiye**

Roles: Data Curation, Formal Analysis, Validation

**Trudy Mhlanga**

Roles: Project Administration, Conceptualization, Writing – Original Draft Preparation

**Efison Dhodho**

Roles: Validation

**Tinashe Chinyanga**

Roles: Funding Acquisition, Supervision

